# Large-scale evaluation of outcomes following a genetic diagnosis in children with severe developmental disorders

**DOI:** 10.1101/2023.10.18.23297202

**Authors:** Harriet Copeland, Karen J Low, Sarah Wynn, Ayesha Ahmed, Victoria Arthur, Meena Balasubramanian, Katya Bennett, Jonathan Berg, Marta Bertoli, Lisa Bryson, Catrin Bucknall, Jamie Campbell, Kate Chandler, Jaynee Chauhan, Amy Clarkson, Rachel Coles, Hector Conti, Philandra Costello, Tessa Coupar, Amy Craig, John Dean, Amy Dillon, Abhijit Dixit, Kathryn Drew, Jacqueline Eason, Francesca Forzano, Nicky Foulds, Alice Gardham, Neeti Ghali, Andrew Green, William Hanna, Rachel Harrison, Mairead Hegarty, Jenny Higgs, Muriel Holder, Rachel Irving, Vani Jain, Katie Johnson, Rachel Jolley, Wendy Jones, Gabriela Jones, Shelagh Joss, Ruta Kalinauskiene, Farah Kanini, Karl Kavanagh, Mahmudur Khan, Naz Khan, Emma Kivuva, Nayana Lahiri, Neeta Lakhani, Anne Lampe, Sally Ann Lynch, Sahar Mansour, Alice Marsden, Hannah Massey, Shane McKee, Shehla Mohammed, Swati Naik, Mithushanaa Nesarajah, Ruth Newbury-Ecob, Fiona Osborne, Michael J Parker, Jenny Patterson, Caroline Pottinger, Matina Prapa, Katrina Prescott, Shauna Quinn, Jessica A Radley, Sarah Robart, Alison Ross, Giulia Rosti, Francis Sansbury, Ajoy Sarkar, Claire Searle, Nora Shannon, Debbie Shears, Sarah Smithson, Helen Stewart, Mohnish Suri, Shereen Tadros, Rachel Theobald, Rhian Thomas, Olga Tsoulaki, Pradeep Vasudevan, Maribel Verdesoto, Emma Vittery, Sinead Whyte, Emily Woods, Thomas Wright, David Zocche, Helen V Firth, Caroline F Wright, the DDD Study

## Abstract

**Objective:** We sought to evaluate outcomes for clinical management following a genetic diagnosis from the Deciphering Developmental Disorders (DDD) Study.

**Design:** Individuals in the DDD study who had a pathogenic/likely pathogenic genotype in the DECIPHER database were selected for inclusion (n=5010). Clinical notes from regional clinical genetics services notes were reviewed to assess pre-defined clinical outcomes relating to interventions, prenatal choices, and information provision.

**Results:** Outcomes were recorded for 4237 diagnosed probands (85% of those eligible) from all 24 recruiting centres across the UK and Ireland. Additional diagnostic or screening tests were performed in 903 (21%) probands through referral to a range of different clinical specialties, and stopped or avoided in a further 26 (0.6%). Disease-specific treatment was started in 85 (2%) probands, including seizure-control medications and dietary supplements, and contra-indicated medications were stopped/avoided as no longer necessary in a further 20 (0.5%).

The option of prenatal/preimplantation genetic testing was discussed with 1204 (28%) families, despite the relatively advanced age of the parents at the time of diagnosis.

Importantly, condition-specific information or literature was given to 3214 (76%) families, and 880 (21%) were involved in family support groups. In the most common condition (KBG syndrome; 79 (2%) probands), clinical interventions only partially reflected the temporal development of phenotypes, highlighting the importance of consensus management guidelines and patient support groups.

**Conclusions:** Our results underscore the importance of achieving a clinico-molecular diagnosis to ensure timely onward referral of patients, enabling appropriate care and anticipatory surveillance, and for accessing relevant patient support groups.

## INTRODUCTION

Despite widespread use of genomic testing in children with developmental disorders (DD), relatively little has been documented about the outcomes following a genetic diagnosis in this group of patients[1]. Steady advances in genomic technologies, including DNA microarray analysis and exome/genome sequencing, have resulted in the identification of a monogenic cause in around half of individuals affected with a presumed genetic DD[2–4]. The value of a diagnosis to the family has been well documented (https://www.undiagnosed.org.uk/support-information/what-does-getting-a-genetic-diagnosis-mean/), including genetic counselling, accessing patient support groups and reproductive planning[5]. However, the value for clinical management has been less clearly documented, and it has sometimes been assumed that in many cases nothing different can be done to manage the affected child[6], rendering a precise molecular diagnosis an additional detail rather than a pivotal point in the ongoing management of the child and their family.

We sought to investigate outcomes in families affected by severe DD where a genetic diagnosis was made through the Deciphering Developmental Disorders (DDD) Study. By including data from across the whole of the UK and Republic of Ireland, we were able to systematically analyse interventions in >4,200 diagnosed probands and evaluate management of individuals affected by the same syndromes.

## METHODS

### Eligibility

Probands with severe previously undiagnosed developmental disorders were recruited into the DDD study and analysed using microarrays and exome sequencing, as described previously[7–9]. Probands were selected for follow-up to investigate outcomes if they had received a likely diagnosis from the DDD study reported to referring clinical geneticists via DECIPHER[10] as of 8 March 2021 (n=5010), herein defined as a clinician-annotated pathogenic/likely pathogenic genotype[4], or *de novo* mutation or biallelic loss-of-function variant in a DDG2P gene[11].

### Data collection

Parental ages, quantitative growth data and Human Phenotype Ontology (HPO) terms were prospectively collected on all probands in the DDD study[12]. A clinical outcomes questionnaire was subsequently designed based on a pilot study[1], including questions relating to treatment, testing/screening, reproductive choice, information provision and adverse outcomes relating to receiving a diagnosis (**Table 1**). In addition to single response questions, information was collected in free-text format on medical interventions (treatments and testing/screening) and adverse outcomes. The questionnaire was codified into a standardised *pro forma* and circulated to each Regional Genetics Service to complete for their diagnosed DDD families using clinical notes from regional clinical genetics services, including a pseudonymized DECIPHER ID linked to the diagnosis for each proband. Data were collated from March 2021 – July 2022. Variants were confirmed in an NHS diagnostic laboratory where appropriate.

**Table 1.** Questionnaire results from 4237 diagnosed families in the DDD study.

| Topic | Question | Yes | No | Unknown |
| --- | --- | --- | --- | --- |
| Interventions | Is a diagnosis specific treatment available?* | 144 (3%) | 3928 (93%) | 165 (4%) |
| Interventions | Were any one-off investigations performed as a result of diagnosis?* | 749 (18%) | 3325 (78%) | 163 (4%) |
| Interventions | Was the proband referred to a different specialty for any screening?* | 799 (19%) | 3295 (78%) | 143 (3%) |
| Interventions | Were there any interventions avoided as a result of diagnosis?* | 53 (1%) | 3884 (92%) | 300 (7%) |
| Interventions | If dx earlier could any interventions have been avoided? | 418 (10%) | 3387 (80%) | 432 (10%) |
| Interventions | Additional research/clinical trials available? | 1207 (28%) | 2483 (59%) | 547 (13%) |
| Reproductive | Has there been any pregnancies since the result? | 235 (6%) | 3006 (71%) | 996 (24%) |
| Reproductive | Would PND be an option if family wished and applicable? | 3029 (71%) | 544 (13%) | 664 (16%) |
| Reproductive | Was PND discussed in clinic? | 1205 (28%) | 2432 (57%) | 600 (14%) |
| Reproductive | Was PND performed? | 78 (2%) | 3741 (88%) | 418 (10%) |
| Reproductive | Was PGD discussed in clinic? | 340 (8%) | 3293 (78%) | 604 (14%) |
| Reproductive | Was PGD performed? | 31 (1%) | 3809 (90%) | 397 (9%) |
| Information/support | Is diagnosis specific information available? | 2798 (66%) | 1076 (25%) | 363 (9%) |
| Information/support | Was this information given/signposted to the family? | 2480 (59%) | 1308 (31%) | 449 (11%) |
| Information/support | Were the family given any scientific literature about the condition? | 1563 (37%) | 2189 (52%) | 485 (11%) |
| Information/support | Were the family included in a scientific paper? | 772 (18%) | 2651 (63%) | 814 (19%) |
| Information/support | Has the family been involved in a patient support group? | 880 (21%) | 1395 (33%) | 1962 (46%) |
| Adverse outcomes | Are there any known adverse outcomes?* | 95 (2%) | 3750 (89%) | 392 (9%) |

\* Further information requested in free text form in separate table

## RESULTS

### Overview of cohort

Outcomes data were recorded on 4,237 diagnosed DDD probands (47% female) by 24 Regional Genetics Services across the UK and Republic of Ireland (range = 42-316 per centre, **Figure 1**). Diagnoses included both small single-gene variants and large multigenic structural variants, with inheritance patterns[4] including autosomal dominant (68% *de novo*, 7% inherited from an affected parent, 8% with unknown inheritance), autosomal recessive (11%), X-linked (5%), as well as multiple diagnoses with different inheritance classes (1%). The average time from recruitment to result was 3.8 years (range: 1.1-9.4 years), at which point probands were an average of 12 years old (range: 1.8-55 years) and parents were an average of 44 years old (range: 20-90 years; **Figure 2**).

**Figure 1.**
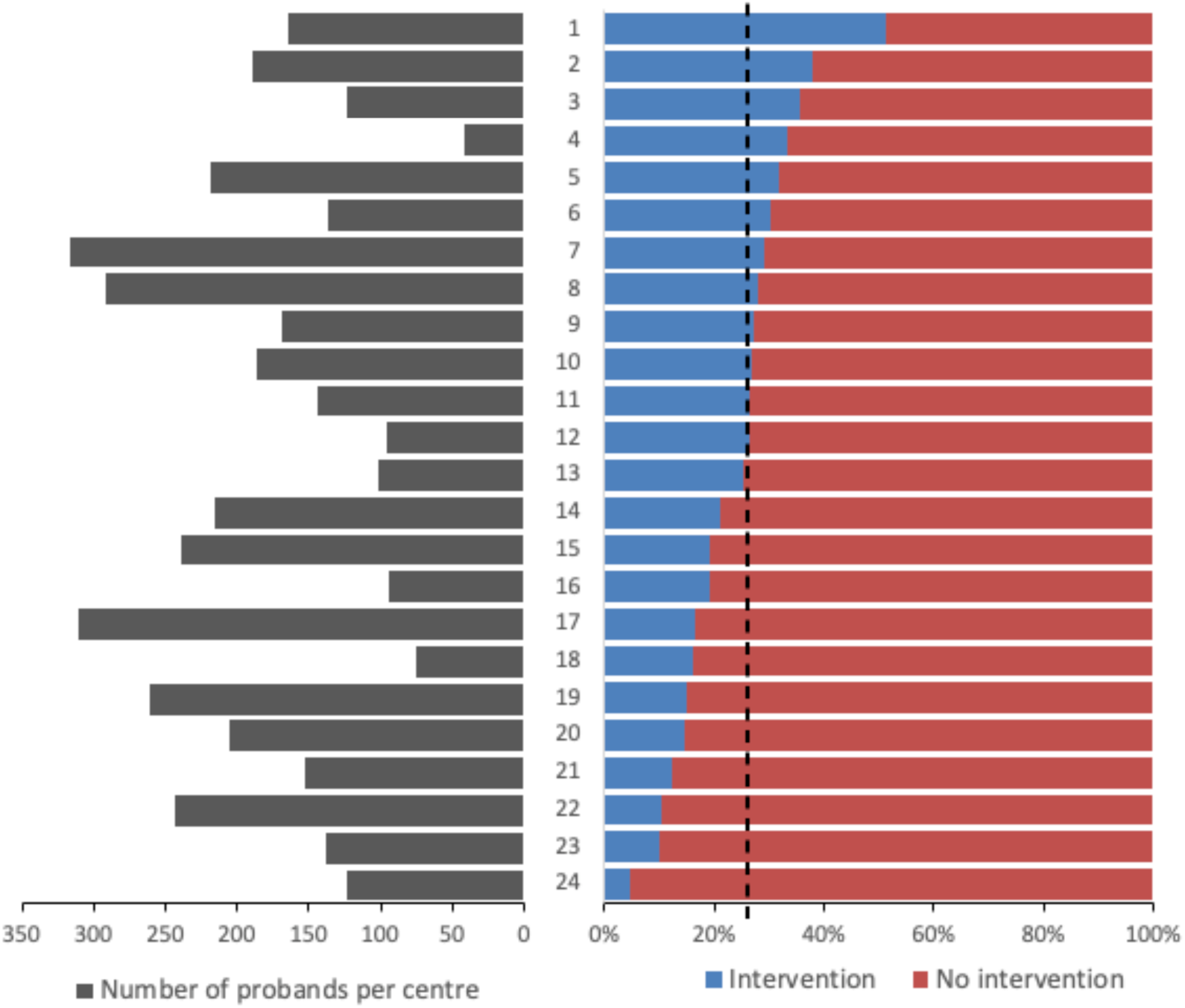
Summary of diagnosed DDD probands per Centre. Number of diagnosed DDD probands included in study (left) and percentage with interventions (treatment/testing; right) separated by the 24 Regional Genetic Services across the UK and Ireland. Black dotted line = average across study.

**Figure 2.**
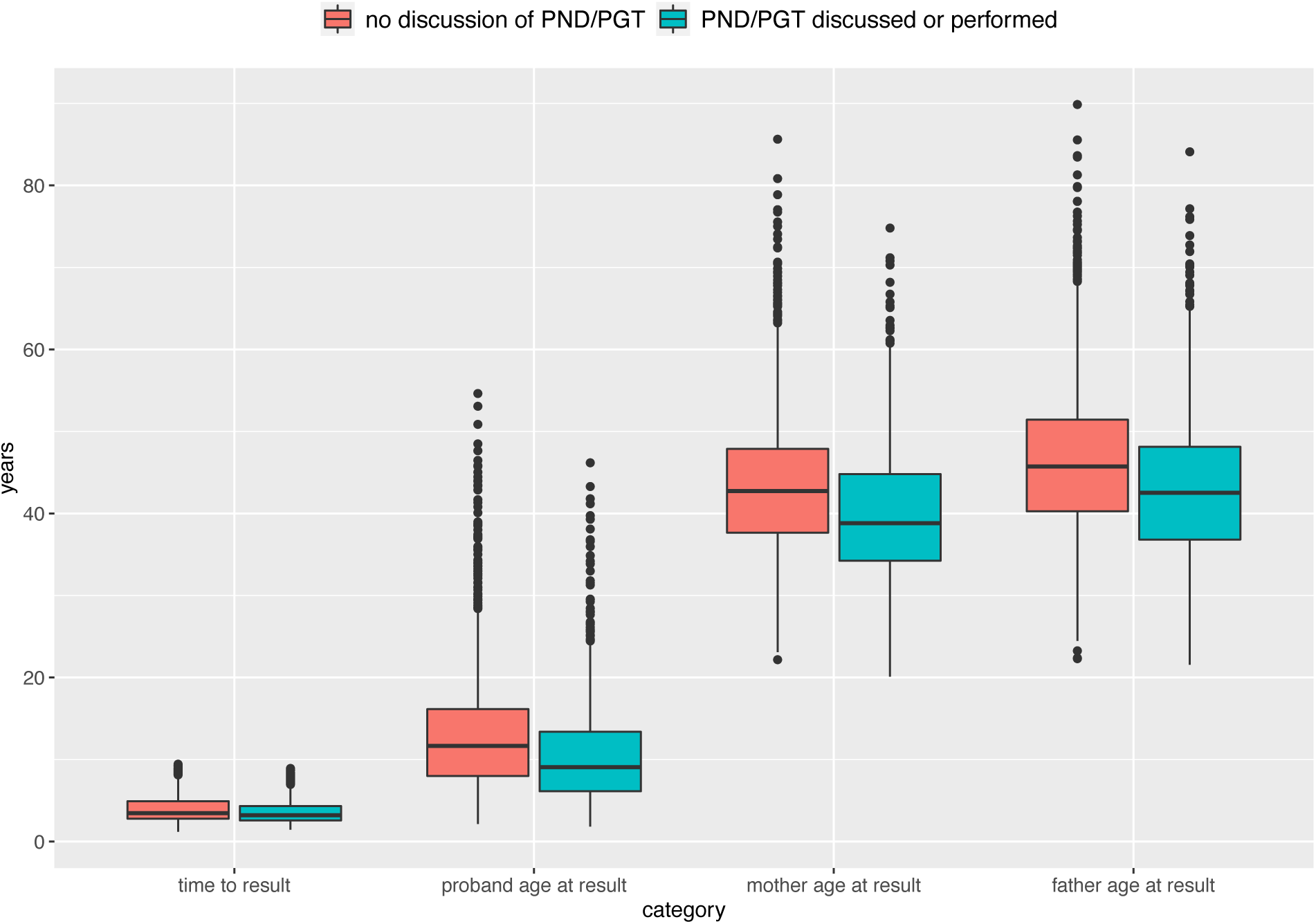
Time to result and age of probands and parents at the point of diagnosis. PND = prenatal diagnosis; PGT = preimplantation genetic testing; red = no record of prenatal testing being discussed with the family; green = prenatal testing discussed or performed.

### Management of proband

An overview of the clinical outcomes that occurred following a diagnosis in 4,237 DDD families are summarised in **Table 1** and **Figure 3**. Importantly, clinical management of the affected individual changed in 24% of diagnosed DDD probands as a result of receiving a genetic diagnosis, which ranged from 5-51% across the different regional genetics services (**Figure 1**). This range partly reflects differences in workforce capacity across different centres. Treatment was altered in 143 probands (3%), which included starting, reviewing, stopping, or avoiding specific therapies. Recurrently prescribed medications included drugs to control seizures (e.g. carbamazepine, clonazepam, lamotrigine, topiramate) and specific dietary supplements (e.g. folate, creatinine, carnitine, ornithine). Interventions include probands who accessed prophylactic treatment to reduce the risk of condition specific complications (e.g. retinal detachment in Stickler syndrome). Further medical investigations were performed in 937 probands (22%) through referral to a wide range of non-genetics specialists for further clinical input to manage associated phenotypes, including screening and/or non-genetic diagnostic testing (**Figure 3**). The largest number of referrals were made to cardiology (28%), followed by nephrology (13%), ophthalmology (11%), radiology (10%), neurology/paediatric neurology (7%), endocrinology (7%), primary care (4%), condition-specific or specialist metabolic clinics (4%), audiology (3%), dentistry (2%), dermatology (2%) and orthopaedics (1%) as well as Ear Nose and Throat (ENT), respiratory, general paediatrics, psychiatry/clinical psychology and urology (all <1%). A third of referred probands were referred for multiple different investigations or to multiple non-genetics specialists to manage different aspects of their phenotype, highlighting the complexity of genetic DD syndromes. Free-text information gathered also indicated that additional phenotypic features were detected and managed in many probands following these referrals, reflecting the value of timely diagnosis and referral for identifying complications and providing appropriate multi-disciplinary care. In 418 probands (10%), it was reported that some interventions (such as MRI scans and muscle biopsies) could have been avoided if the diagnosis had been made earlier.

**Figure 3.**
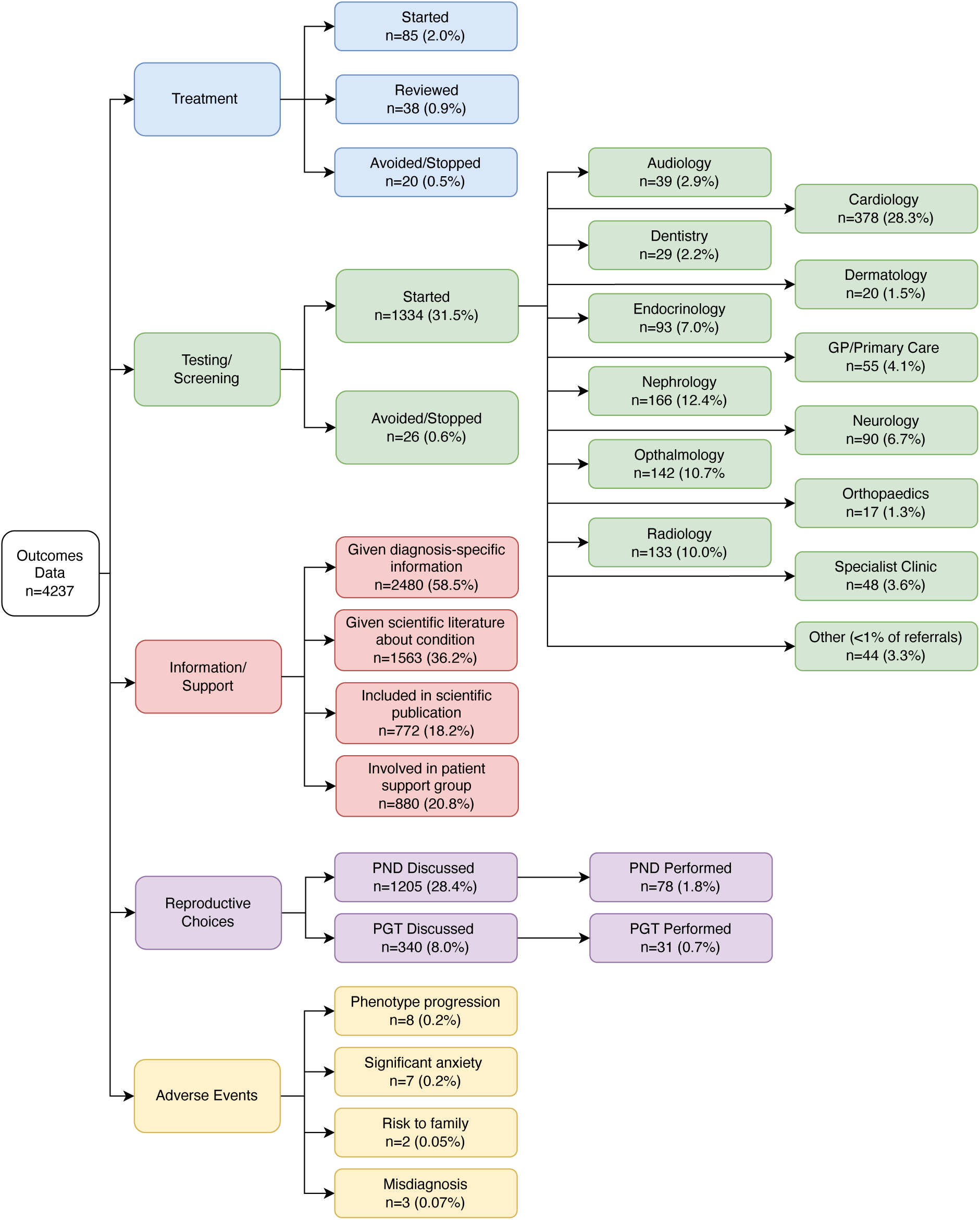
Flowchart summarising outcomes following a genetic diagnosis in the DDD study. PND = prenatal diagnosis; PGT = preimplantation genetic testing.

### Management of family

In addition to medical management of the affected proband, we also investigated wider clinical management of the family following their diagnosis. Condition-specific information or support was provided to 3214 families (76%), including scientific literature and/or patient information leaflets. Remarkably, 772 (18%) of families had been included in condition-specific scientific publications, which likely reflects the rarity and recent discovery of many of the disease-associated genes. At the time of data collection, prenatal diagnosis or preimplantation genetic testing (PND/PGT) had been discussed with 1222 families (29%) and performed in 103 (2%). It is likely these proportions would have been higher had parents been younger at the point of receiving the diagnosis (**Figure 2**), and only 235 (6%) of parents had a confirmed pregnancy since receiving their child’s genetic diagnosis. Finally, reflecting the fact that receiving a diagnosis does not always provide welcome news, a diagnosis-related adverse outcome was reported in 20 families (0.5%), in whom parental or patient anxiety resulted in additional clinic appointments. Reasons given for anxieties related to a range of issues, including the possibility of phenotype progression (based on other individuals affected with the same condition), the prospect of additional interventions, the lack of diagnosis-specific information, potential risks to other family members, and changes to a previous diagnostic result (either a previous missed or mis-diagnosis[13]).

### Data aggregation to build knowledge

We further sought to compare phenotypes and outcomes between probands of different ages diagnosed with the same condition. In our dataset, 37 genes had diagnostic variants in ≥20 probands, together accounting for 1218 (29%) of diagnoses[4]. Of these, we focused on three well-established exemplar genes: *ANKRD11* (KBG syndrome; n=79)[14], which has the largest number of DDD diagnoses; *CTNNB1* (neurodevelopmental disorder with spastic diplegia and visual defects, NEDSDV; n=30)[15], in which there is a clinical imperative for ophthalmic surveillance; and *NSD1* (Sotos syndrome; n=20)[16], in which the highest proportion of DDD probands (65%) had medical interventions following a diagnosis. Using HPO terms and quantitative phenotypes grouped by age and system, we created a quasi-natural history for the conditions and overlaid information about when and how often particular interventions occurred (**Figure 4**).

**Figure 4.**
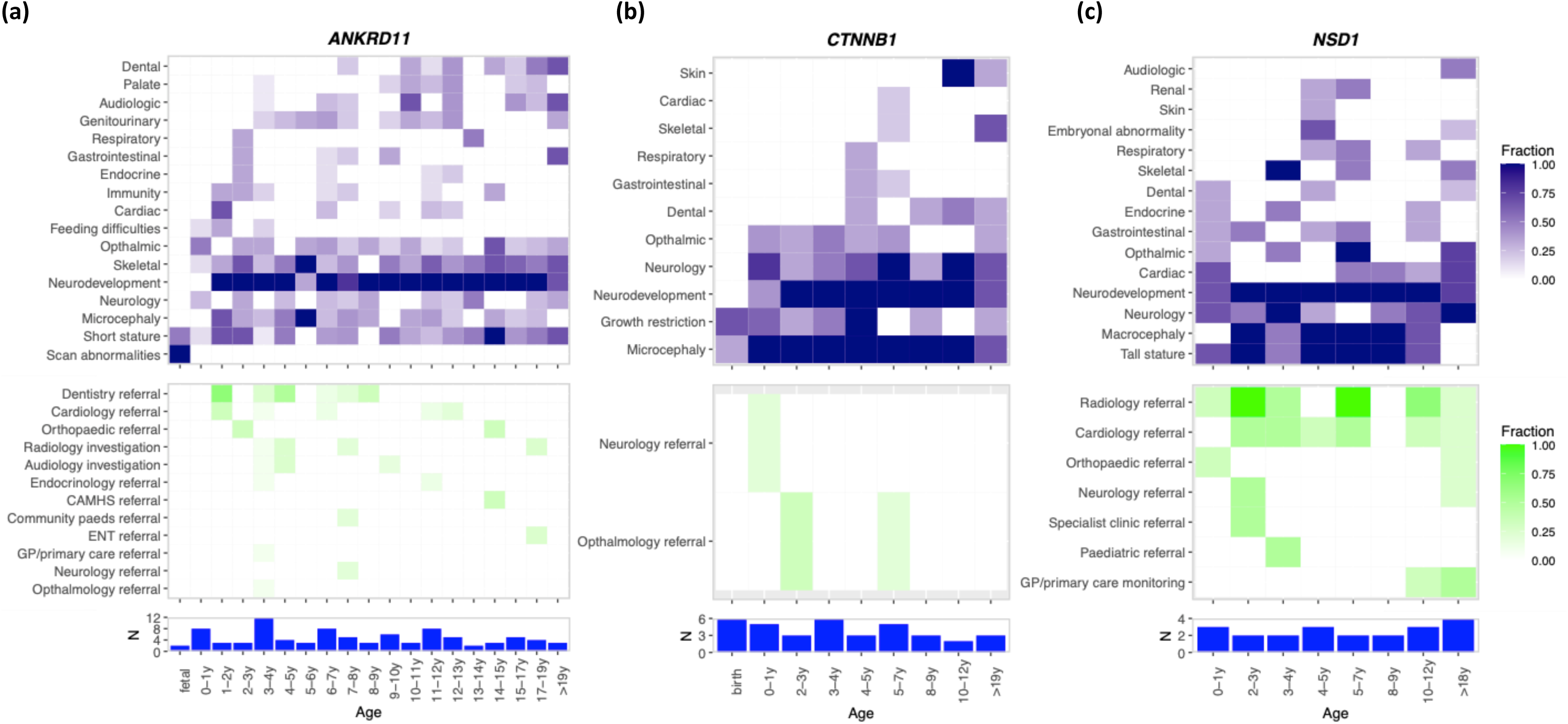
Quasi-natural history of disease and summary of interventions for DDD probands diagnosed with the same condition. **(a)** *ANKRD11,* **(b)** *CTNNB1* and **(c)** *NSD1* (right). Top panel – heatmap of phenotypes grouped by system and age; middle panel – heatmap of interventions grouped by age; bottom panel – histogram of number of probands in each group based on age at recruitment.

For *ANKRD11,* the phenotype heatmap (**Figure 4a**) demonstrates a multi-system disorder with variable expression. Short stature and neurodevelopmental features are strongly consistent throughout the age range but there is an age-dependent emergence to other features such as dental and audiologic phenotypes. The spread of other phenotypes is consistent with the body of literature already available in KBG Syndrome, but demonstrates a highly visual quasi-natural history, useful for both parents and clinicians alike when determining management plans at a point in time. Interventions in *ANKRD11* patients demonstrated large-scale variability across the group, which is likely associated with the timing of the emergence of published clinical recommendations[17]. In contrast, the heatmap for *CTNNB1* (**Figure 4b**) illustrates a more tightly defined range of phenotypic features, demonstrating a severe early onset neurodevelopmental disorder with postnatal onset microcephaly. Interestingly, we did not observe a consistent pattern of ophthalmology referrals amongst these patients, despite a 40% risk of retinal detachment requiring regular eye surveillance to prevent total blindness[18]. This observation is potentially due to variability in data collection for onward referral and the severity of the phenotype precluding referral, but suggests an opportunity to alert clinicians to the need for ophthalmology referral in these patients. By comparison, the well-documented recommendations for baseline investigations and referrals were evident in our data for in *NSD1* (**Figure 4c**), as was the established evolution of the phenotype with age[19]. Interestingly, although patterns of phenotype progression are apparent with increasing age, all three conditions show a degree of variable expressivity, with only a few phenotypes universally present. Clinical interventions across all three conditions appear to be somewhat sporadic, and only partially reflect the temporal development of phenotypes, suggesting that systematic improvements could be made to referral practices to ensure equity of access to the most appropriate care.

### Benefits of support groups

Finally, we found that 880 (20.8%) of diagnosed DDD families were involved in patient support groups. In addition to umbrella patient organisations supporting families with genetic conditions and pre-existing condition-specific organisations, numerous new condition-specific patient support groups were created as a direct result of disease-gene discovery in the DDD Study. These groups range from small parent-led social media (e.g. Facebook) groups that bring patients and families together to share experiences, to the development of registered charities and foundations. We also note that, over the course of the study, DDD clinical collaborators have contributed to authoring >40 single gene patient information leaflets in collaboration with Unique (https://rarechromo.org/disorder-guides/).

## DISCUSSION

We have retrospectively recorded and analysed outcomes following a genetic diagnosis in 4237 families in the DDD study. We have shown that around a quarter of individuals affected by a severe DD received a change in medical management following their genetic diagnosis, primarily through a range of referrals to non-genetics specialties for additional testing and surveillance. The clinical impact of a precise molecular diagnosis on the management pathway for an individual patient thus enables a precision medicine approach and the provision of appropriate care, sometimes preventing particular phenotypes from developing. The likely increased demand for specialist assessments following a genetic diagnosis also needs to be costed and provided. Additionally, at least three-quarters of families were given condition-specific information, which supports understanding and family adaption to a genetic diagnosis. Very few adverse outcomes were reported, suggesting that the anxiety and other mental health implications associated with receiving genetic results from a large genomics research study delivered via an expert clinical service were generally low.

We have also presented a novel approach to creating a quasi-natural history of specific genetic conditions, using data from multiple affected probands of different ages. The richness of phenotype data in KBG syndrome in particular shows the variable expressivity of this highly penetrant condition, and highlights when and how likely particular phenotypes are to manifest. However, the link between the emergence of clinical phenotypes and the necessary clinical interventions is weak, and may vary both within a condition and between services. We hope that these representations of phenotypes and intervention data with age will provide better prognostic information to clinicians and patients, and catalyse the development of consensus management guidelines. In addition, the growing size and number of disorder-specific family support groups should be recognised and welcomed by both the clinical and patient communities, and may provide a mechanism by which referral and clinical management practices could be compared and optimised. Support groups play a vital role in the provision of information and act as a forum for patients and families to share experiences and seek advice from people in a similar situation[20]. Parents and carers of children with DD are at risk of social isolation and emotional distress, which can be exacerbated when the condition is rare[21,22]. Many participants of support groups report positive outcomes, such as reduced isolation and anxiety, improvements in coping skills and increased self-esteem and empowerment[23]. Internet-based support groups also mean that geographical location is no barrier to accessing support and making connections with others[24]. Ultimately, bringing together patients, clinicians and researchers with a common focus on a specific condition can stimulate research, enabling co-development of research questions and providing a vehicle for both recruitment and dissemination of findings.

This large-scale, nation-wide study was made possible through an extensive network of regional clinical collaborators across the National Health Service and Health System in the UK and Ireland. However, there are significant challenges to gathering comparable data on thousands of families under the care of hundreds of clinicians spread across 24 different sites.

Due to the large size and geographical spread of the study, we did not attempt to gather information directly from parents or probands relating to social, educational or other non-clinical outcomes, though there is little doubt that receiving a formal diagnosis can be of immense value to families. Provision of social, financial and educational support should be based on an individual’s need, but families often report that a diagnostic label can be extremely helpful when advocating for their child’s needs[1,25,26]. Within each clinic, individual data collectors were limited to information available in their local genetics notes, where the level of detail routinely recorded can vary substantially – exacerbated by the move from paper towards electronic health records – hampering our ability to compare findings between services. Moreover, the size and expertise of data collection teams varied across the sites, potentially resulting in different ways of reporting similar outcomes. There may also be differences between clinicians and regions in referral practices (e.g. refer versus test onsite) as well as the timing and purpose of testing (e.g. diagnostic versus screening, etc).

We were also limited by the retrospective collection of outcomes data, recorded at a single point in time but relating to diagnoses returned over the course of a 7-year period. This approach cannot account for the development of clinical guidelines and dissemination of best practice over time. This issue is exemplified by KBG Syndrome, for which clinical management recommendations were published in 2016, after most *ANKRD11* diagnoses were returned in DDD[17]. Similarly, we were limited by the prospective collection of phenotypes at recruitment, which does not take account of phenotypic progression. It was not always possible to determine whether a particular clinical action resulted directly from the genetic diagnosis or from the appearance of a phenotype. Our results are skewed both by the high proportion of diagnostic *de novo* variants and the relatively advanced age of parents at the point of receiving a diagnosis, which may have reduced the appropriateness of reproductive counselling and limited parental opportunity for further testing. Finally, even within our large dataset, due to the rareness of individual conditions, there were relatively small numbers of probands with the same conditions, which reduced our ability to create accurate quasi-natural histories across different age groups. Ideally, longitudinal phenotype collection on individuals would enable true natural histories to be collected and compared, and the aggregation of data on larger numbers of patients through databases such as DECIPHER will enable these data to be systematically analysed and widely shared.

In conclusion, we have demonstrated that it is both possible and useful to collect outcomes data from clinical genetics services on the impact of receiving a genetic diagnosis. Making an accurate genetic diagnosis is often crucial for directing clinical management of affected individuals and providing advice regarding risks to other family members including reproductive advice. Although molecularly-targeted treatments for monogenic DDs are still limited, more will no doubt become available as new technologies develop. Our findings highlight the importance of onwards referral to ensure the best care for patients and families affected by rare diseases, and also underscore the value of developing best practice guidelines to ensure equity of access to appropriate clinical interventions.

## Data Availability

All data produced in the present study are available upon reasonable request to the authors

https://www.deciphergenomics.org/

## ACKNOWLEDGEMENTS

We thank Professors Michael Parker and Matthew Hurles for their ongoing involvement and leadership of the DDD study. The authors are deeply indebted to the patients and families involved in the DDD study, as well as the UK NHS and Irish HSE genetics services. The DDD study presents independent research commissioned by the Health Innovation Challenge Fund [grant number HICF-1009-003], a parallel funding partnership between the Wellcome Trust and the Department of Health, and the Wellcome Sanger Institute [grant number WT098051].

The study has UK Research Ethics Committee approval (10/H0305/83, granted by the Cambridge South REC, and GEN/284/12 granted by the Republic of Ireland REC). This study was supported by the National Institute for Health and Care Research Exeter Biomedical Research Centre. The research team acknowledges the support of the National Institute for Health Research, through the Comprehensive Clinical Research Network. DECIPHER is funded by Wellcome [WT223718/Z/21/Z]. This research was funded in whole or in part by the Wellcome Trust. For the purpose of open access, the author has applied a CC-BY public copyright licence to any author accepted manuscript version arising from this submission. The views expressed are those of the author(s) and not necessarily those of the Wellcome, NIHR or the Department of Health and Social Care. This study was supported by the National Institute for Health and Care Research Exeter Biomedical Research Centre. The views expressed are those of the author(s) and not necessarily those of the NIHR or the Department of Health and Social Care.

